# Cost-effectiveness of HPV-based versus VIA-based cervical cancer screening among women living with HIV in Masaka District, Uganda

**DOI:** 10.64898/2026.07.09.26357621

**Authors:** Asiimwe Anita Yvonne, Bongole Godfrey, Namuhani Noel

**Affiliations:** Department of Health Policy Planning and Management, School of Public Health, Makerere University, Kampala, Uganda

## Abstract

Cervical cancer is a leading cause of cancer-related mortality among women in Uganda, with women living with HIV (WLHIV) at disproportionate risk due to immunosuppression-driven persistence of human papillomavirus (HPV). Despite national guidelines recommending HPV testing as the preferred screening modality, resource constraints drive continued reliance on visual inspection with acetic acid (VIA), and locally generated cost-effectiveness evidence for WLHIV is limited. This study evaluated the cost-effectiveness of HPV-based versus VIA-based cervical cancer screening among WLHIV in Masaka District, Uganda.

A provider-perspective cost-effectiveness analysis was conducted using data from January to December 2024. Costs were estimated using an ingredient-based micro-costing approach capturing personnel, consumables, equipment, and overheads. A total of 1,732 WLHIV aged 25–65 years attended Uganda Cares Masaka: 1,404 screened by HPV-based testing and 326 by VIA. A decision-tree model simulated screening pathways, costs, and outcomes. The primary effectiveness measure was the number of positive cases detected and treated; the incremental cost-effectiveness ratio (ICER) was the primary economic outcome. Deterministic one-way and probabilistic (1,000 Monte Carlo iterations) sensitivity analyses were conducted.

HPV-based screening detected 448 positives from 1,404 women screened (31.9%) versus 54 from 326 (16.6%) under VIA. The cost per woman screened was USD 2.58 (HPV) and USD 1.78 (VIA). The ICER was USD 2,895 per additional positive case detected, within the range of ICER estimates reported for cost-effective cervical cancer screening in comparable low- and middle-income settings. The ICER was most sensitive to HPV test kit costs and VIA overhead costs. All 1,000 probabilistic simulations remained in the northeast quadrant of the cost-effectiveness plane, confirming robustness.

HPV-based screening is more effective and cost-effective than VIA for cervical cancer screening among WLHIV in Masaka District. These findings support national scale-up of HPV testing within integrated HIV care settings, contingent on procurement efficiencies and strengthened laboratory infrastructure.

## Introduction

Cervical cancer ranks as the fourth most common cancer in women globally, with an estimated 604,000 new cases and 342,000 deaths recorded in 2020 [1]. The burden falls overwhelmingly on low- and middle-income countries (LMICs), which account for approximately 90% of global cervical cancer mortality [2]. Sub-Saharan Africa bears the greatest regional burden, with Uganda recording one of the highest incidence rates in the world at approximately 54.8 to 56.2 per 100,000 women and an estimated 6,959 new cases and 4,607 deaths annually [3, 4]. Cervical cancer is the second most common cancer among Ugandan women, and 80% of cases are diagnosed at advanced stages when treatment options are limited and survival is poor [4].

Women living with HIV (WLHIV) face a substantially elevated risk of cervical cancer compared with the general population. Immunosuppression arising from HIV infection accelerates the persistence of high-risk human papillomavirus (HPV) and the progression from cervical intraepithelial neoplasia to invasive cancer [5, 6]. Systematic reviews indicate that WLHIV are up to six times more likely to develop cervical cancer than HIV-negative women [7]. In Uganda, HIV prevalence among women stands at 6.4%, higher than the national average of 4.9%, and Masaka District records an HIV prevalence of 9.8% to 11.7% among adults aged 15 to 49 years [8, 9], making it one of the highest-burden districts in the country. This dual burden of HIV and HPV creates an urgent imperative for targeted and cost-efficient cervical cancer screening strategies in this population.

Two cervical cancer screening strategies are actively deployed at the primary health care level in Uganda. Visual inspection with acetic acid (VIA) involves applying 3 to 5% acetic acid to the cervix; abnormal cells turn white within one to two minutes, enabling immediate visual diagnosis and same-day treatment in a screen-and-treat model [10]. VIA is low-cost, requires minimal laboratory infrastructure, and can be performed by trained non-physician health workers, making it well-suited to resource-limited settings [11]. However, VIA has relatively low and operator-dependent sensitivity, raising concerns about missed diagnoses and downstream costs associated with treating advanced-stage disease [12]. HPV-based screening uses molecular assays, including the GeneXpert platform widely deployed in Uganda, to detect DNA from high-risk HPV types in clinician-collected or self-collected cervical specimens [13]. HPV testing offers substantially higher and more standardised sensitivity than VIA [11, 12], and the Uganda National Cervical Cancer Screening and Treatment Guidelines (2023) now recommend it as the primary screening modality, particularly for WLHIV [13].

Despite this policy direction, decisions about deploying and scaling HPV testing have not been sufficiently supported by locally generated cost-effectiveness evidence specific to WLHIV. Economic evaluations from Burkina Faso and Malawi provide relevant regional comparators [14, 15], but may not adequately reflect Uganda’s cost structures, health system constraints, and epidemiological profile. Understanding the cost-effectiveness of HPV testing will inform efforts by policy makers and program implementers to scale up HPV screening among WLHIV in Uganda and other similar settings. This study aimed to evaluate the cost-effectiveness of HPV-based screening compared to VIA-based screening for cervical cancer among WLHIV in Masaka District, Uganda.

## Materials and methods

### Study design

This was a cost-effectiveness analysis using a decision analytic modelling framework. Outcome data were obtained retrospectively from routinely collected facility service delivery records, while cost data were collected prospectively through primary key informant interviews and validated against facility financial records. Reporting followed the Consolidated Health Economic Evaluation Reporting Standards (CHEERS) 2022 checklist [16].

### Study setting

The study was conducted at Uganda Cares Masaka, a key non-governmental healthcare provider delivering HIV care and cervical cancer screening services within the Masaka Regional Referral Hospital campus, Masaka District, Central Uganda. Masaka District has an HIV prevalence of 9.8% among adults aged 15 to 49 years, substantially higher than the national average [8]. Uganda Cares Masaka operates an integrated cervical cancer clinic that provides both HPV-based and VIA-based screening within an established HIV care programme, utilising a screen-and-treat model for VIA-positive findings and GeneXpert-based HPV DNA testing for HPV-based screening.

### Study population

The study population comprised WLHIV aged 25 to 65 years who attended Uganda Cares Masaka for cervical cancer screening between January and December 2024. This age range aligns with national and WHO screening guidelines for WLHIV [13, 17]. Records were included using consecutive sampling of all WLHIV who received screening services during the study period.

### Screening strategies compared

Two strategies were compared. The first was HPV-based screening using GeneXpert molecular assays to detect high-risk HPV strains (HPV-16 and HPV-18). A positive result triggered colposcopy referral and precancerous lesion treatment. The second was VIA-based screening in which 3 to 5% acetic acid was applied to the cervix by trained nurses; acetowhite lesions were recorded as positive findings and eligible for immediate treatment. Both strategies were delivered as part of routine programme care.

### Perspective and time horizon

A provider perspective was adopted, capturing all direct costs borne by the healthcare provider. Patient-borne costs, including transport and time, were excluded. The time horizon was one year (January to December 2024), corresponding to the period of available service delivery data. Discounting was not applied given the one-year horizon [18].

### Data sources

Cost data were collected through structured key informant interviews with nursing staff, laboratory technicians, programme managers, and facility accountants from Uganda Cares Masaka, cross-referenced and validated against procurement invoices, payroll records, utility bills, and administrative expenditure reports. Outcome data were extracted from facility screening registers and electronic records using a standardized data extraction form. Both HPV-based and VIA-based screening were delivered within the same facility (Uganda Cares Masaka) during the same calendar year, ensuring that differences in outcomes and costs between the two arms reflect differences in screening strategy rather than differences in facility capacity, patient population, or care quality. Women were offered both methods during routine counselling and self-selected or were guided to a method based on clinical assessment, which supports comparability of the arms while acknowledging that self-selection may introduce some differential risk profile.

### Costing approach

Costs were estimated using an ingredient-based micro-costing approach incorporating time-driven activity-based costing for personnel time. Cost categories included: (i) direct medical costs, comprising HPV test kits and VIA consumables including acetic acid and speculums; (ii) personnel costs, based on staff time allocated per screening session for nurses and laboratory technicians; (iii) equipment costs, including annualized depreciation of the GeneXpert machine at 2% per annum and clinic equipment; (iv) overhead costs, including administrative expenditure, electricity, water, and building space allocated proportionally by screening volume. All costs were initially captured in Ugandan Shillings (UGX) and converted to United States Dollars (USD) using the Bank of Uganda 2024 average exchange rate of 1 USD = 3,665 UGX [19]. Shared resources were allocated proportionally based on the relative volume of HPV-screened versus VIA-screened women.

### Outcome measures

The primary effectiveness measure was the number of positive cases detected and treated, expressed as an effectiveness probability from the decision-tree model. This was defined as the product of the probability of a positive test result and the probability of treatment among those testing positive. The ICER was the primary economic measure, representing the additional cost per additional positive case detected and treated. Secondary effectiveness measures included positivity rates and treatment rates for each strategy.

### Decision analytic model

A decision-tree model was constructed to simulate screening pathways and their associated costs and outcomes. Two branches corresponded to the two screening strategies. Within each branch, women could receive a positive or negative test result; positive cases were further branched into treated and untreated categories. The model was populated with probabilities derived from facility records and literature-sourced estimates for test sensitivity and specificity [20]. The decision-tree figure will be inserted at Fig 1.

**Fig 1.**
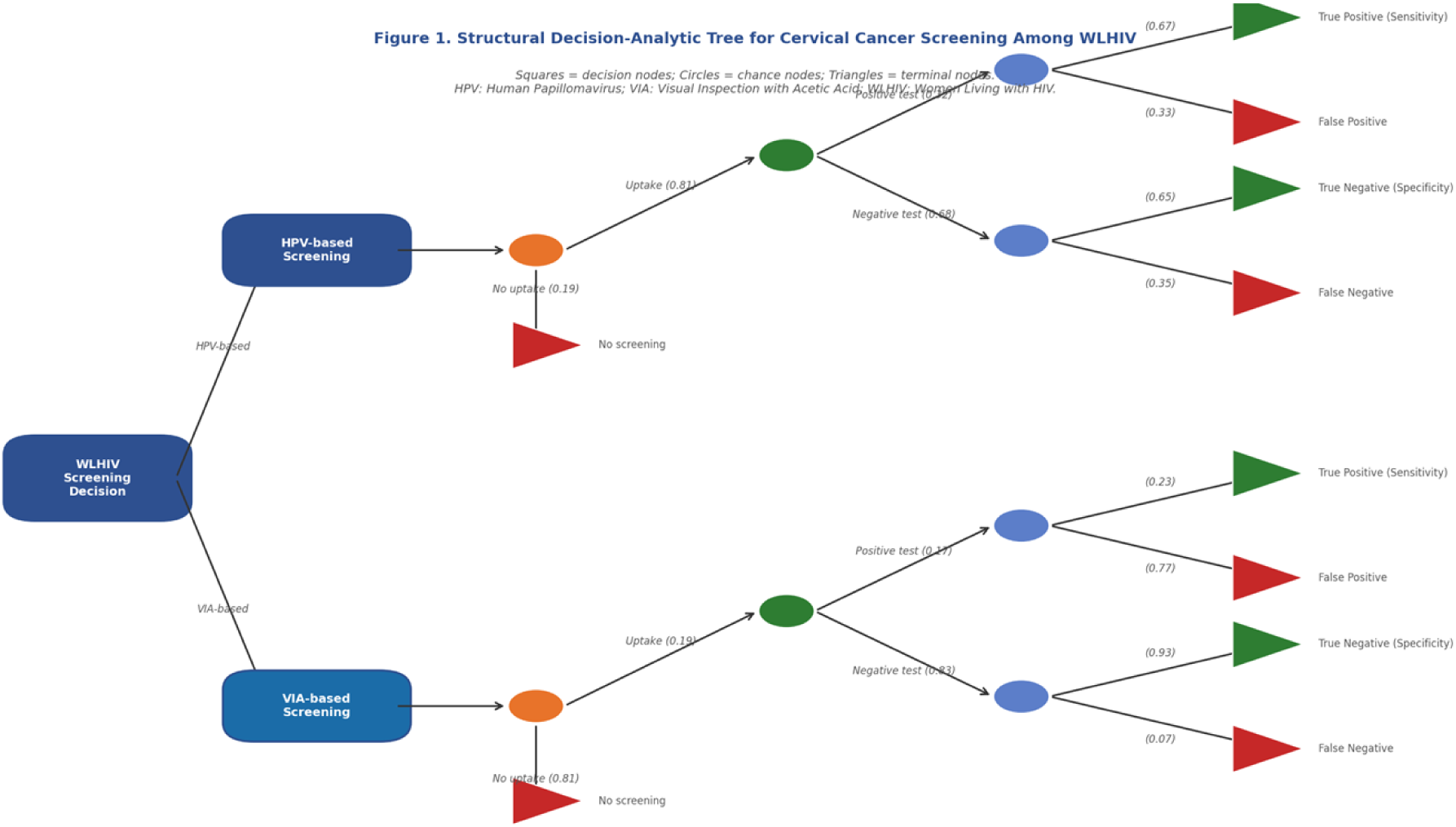
Decision tree model for cervical cancer screening strategies among women living with HIV.

### Cost-effectiveness analysis

The incremental cost-effectiveness ratio (ICER) was calculated using the formula:

ICER = (Cost□□ᵥ - Costᵝᴵᴴ) / (Effectiveness□□ᵥ - Effectivenessᵝᴵᴴ)

where costs represent the total expected costs from the decision-tree model and effectiveness represents the expected effectiveness probabilities. HPV-based screening was assigned as the more costly and more effective strategy. Given that the primary outcome is an intermediate measure (positive cases detected rather than quality-adjusted life years), the ICER was interpreted in the context of published ICER ranges from comparable cervical cancer screening studies in low- and middle-income countries rather than a formal gross domestic product per capita threshold.

### Sensitivity analysis

Two sensitivity analyses were conducted. First, a one-way deterministic sensitivity analysis was performed by varying each key model parameter individually by plus or minus 20% of its base-case value, with results presented as a tornado diagram (Fig 2). Second, a probabilistic sensitivity analysis (PSA) using 1,000 Monte Carlo iterations was conducted using @RISK version 8.2 (Palisade Corporation). Gamma distributions were assigned to cost parameters, beta distributions to probability and proportion parameters, and normal distributions to effectiveness parameters. PSA results are presented on a cost-effectiveness plane (Fig 3).

**Fig 2.**
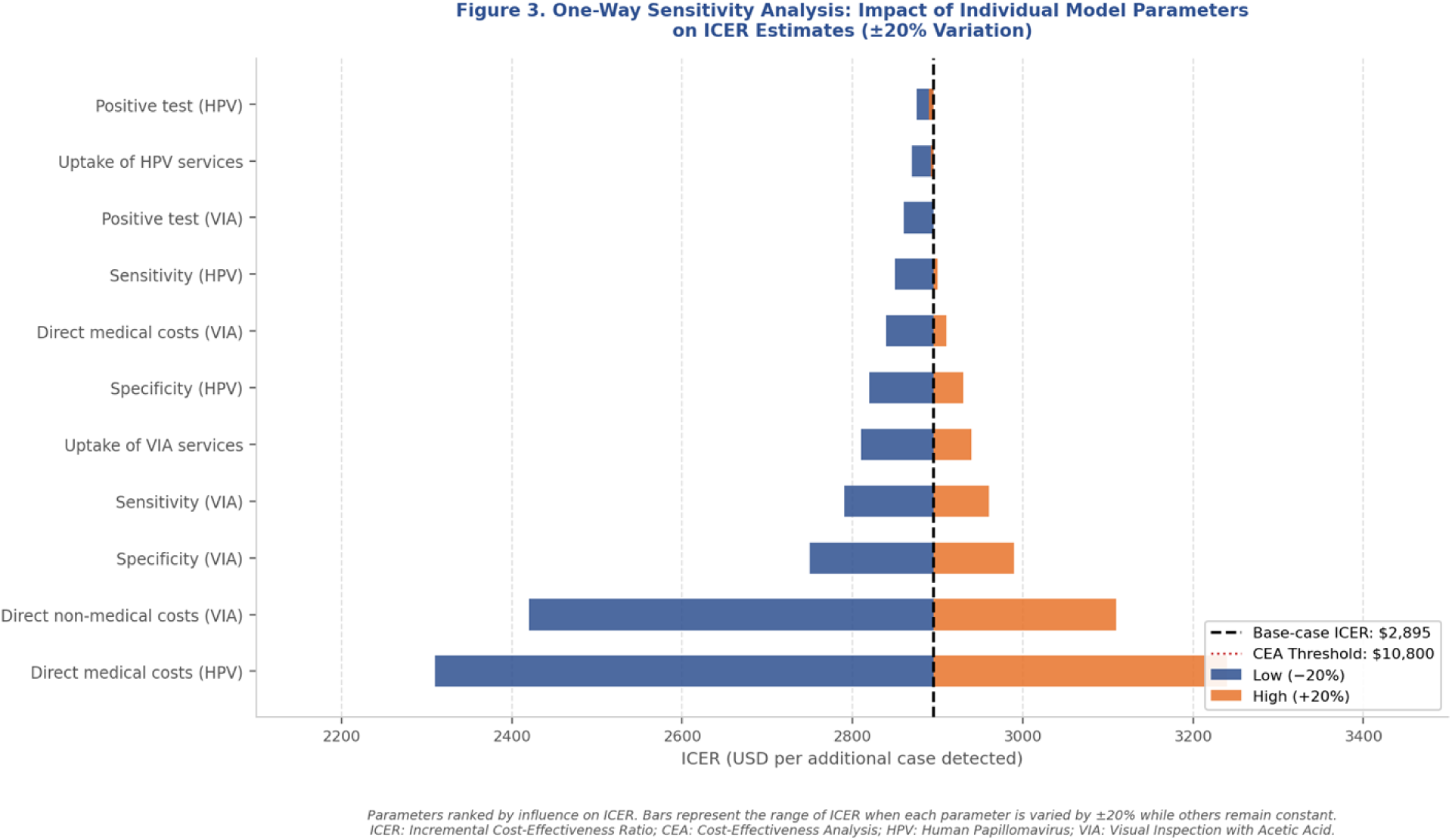
Tornado diagram showing the impact of individual model parameters on the incremental cost-effectiveness ratio.

**Fig 3.**
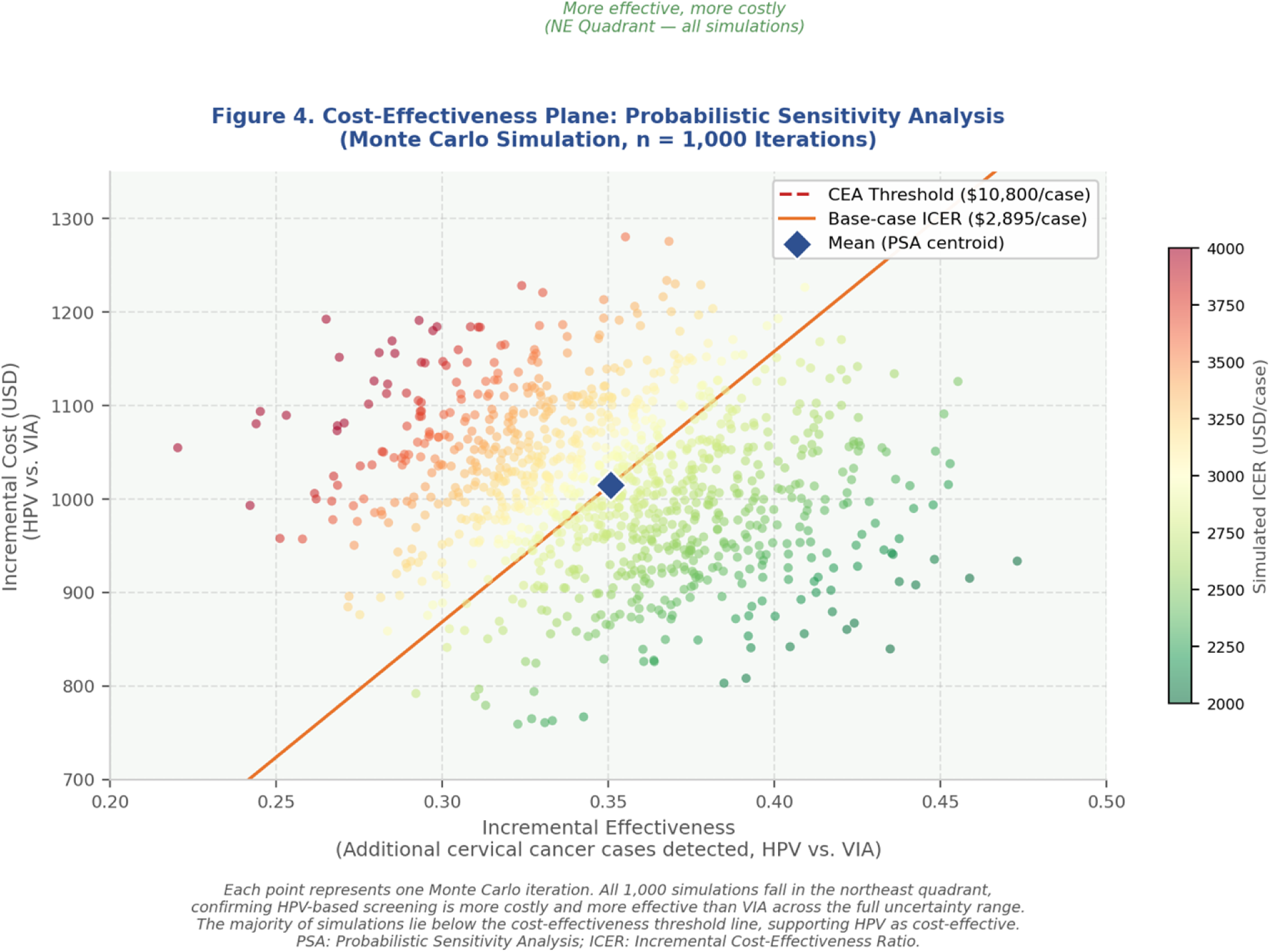
Cost-effectiveness plane from probabilistic sensitivity analysis (1,000 Monte Carlo iterations).

### Statistical and economic analysis software

Statistical calculations were performed in Microsoft Excel 365. Decision-tree modelling and one-way sensitivity analysis were conducted in Microsoft Excel using data tables. Probabilistic sensitivity analysis was performed using @RISK version 8.2 (Palisade Corporation, Ithaca, NY, USA) as a Microsoft Excel add-in.

### Ethical considerations

The study was approved by the Makerere University School of Public Health Research and Ethics Committee. A waiver of informed consent was granted on the grounds that only routinely collected, de-identified, aggregate service delivery data were used. Institutional permission to access records was obtained from Uganda Cares Masaka.

## Results

### Characteristics of the screened population

A total of 1,732 WLHIV attended Uganda Cares Masaka for cervical cancer screening between January and December 2024. Of these, 1,404 (81.1%) were screened using HPV-based testing and 326 (18.9%) using VIA-based screening. The screened population was predominantly aged 25 to 49 years. Detailed participant characteristics by screening strategy are presented in Table 1.

**Table 1.** Characteristics of women screened by strategy.

| Characteristic | HPV-based screening<br>(n = 1,404) | VIA-based screening<br>(n = 326) |
| --- | --- | --- |
| Age range (years) | 25–65 | 25–65 |
| Recommended screening age group, n (%) | 1,404 (100) | 326 (100) |
| Screened, n | 1,404 | 326 |
| Positive cases detected, n (%) | 448 (31.9) | 54 (16.6) |
| Cases treated, n (%) | 420 (93.6) | 53 (98.1) |

### Screening outcomes by strategy

HPV-based screening detected 448 positive cases from 1,404 women screened, yielding a positivity rate of 31.9%. Of those testing positive, 420 (93.6%) received treatment, giving a treatment rate of 93.6%. VIA-based screening detected 54 positive cases from 326 women screened (16.6% positivity rate), with 53 (98.1%) treated. The absolute number of cases treated was substantially higher under the HPV arm (420 versus 53). HPV-based screening demonstrated a sensitivity of 67% and a specificity of 65%, compared with 23% sensitivity and 93% specificity for VIA-based screening. Screening outcomes are summarised in Table 2.

**Table 2.** Screening and treatment outcomes by strategy. HPV, human papillomavirus; VIA, visual inspection with acetic acid.

| Outcome | HPV screened | HPV positives | HPV treated | VIA screened | VIA positives |
| --- | --- | --- | --- | --- | --- |
| Women screened, n | 1,404 | 448 | 420 | 326 | 54 |
| Positivity/treatment rate, % |  | 31.9 | 93.6 |  | 16.6 |
| Sensitivity, % | 67 |  |  | 23 |  |
| Specificity, % | 65 |  |  | 93 |  |

### Cost estimates

Total screening costs were USD 3,636.20 for the HPV arm (1,404 women) and USD 1,778.50 for the VIA arm (326 women). The cost per woman screened was USD 2.58 for HPV-based screening and USD 1.78 for VIA-based screening. Despite the higher per-woman screening cost, the cost per positive case detected was lower under HPV (USD 8.54) compared with VIA (USD 13.09), owing to the substantially higher positivity rate. The largest cost drivers in the HPV arm were HPV test kits (USD 985.19), personnel costs (USD 1,133.80), and administrative overheads (USD 675.60). In the VIA arm, the largest cost contributors were vaginal speculums (USD 746.00) and personnel costs (USD 526.60). Detailed cost parameters are presented in Table 3 and summarized in Table 4.

**Table 3.** Cost parameter breakdown by screening strategy (USD). All costs converted from Ugandan Shillings using the Bank of Uganda 2024 average exchange rate of 1 USD = 3,665 UGX [19]. Shared resources were allocated proportionally by screening volume. USD, United States Dollars; UGX, Ugandan Shillings; HPV, human papillomavirus; VIA, visual inspection with acetic acid.

| Cost item | Unit cost (USD) | HPV quantity | HPV total (USD) | VIA quantity | VIA total (USD) |
| --- | --- | --- | --- | --- | --- |
| HPV test kits | 0.70 | 1,404 | 985.19 | 0 | 0 |
| VIA acetic acid | 0.075 | 0 | 0 | 326 | 24.54 |
| Vaginal speculum | 2.20 (disposable) | 0 | 0 | 326 | 746.00 |
| Electricity | 1.64/day | 1,404 | 49.31 | 326 | 11.45 |
| Water | 0.55/day | 1,404 | 16.44 | 326 | 3.82 |
| Medical sundries | 13.64/day | 1,404 | 411.00 | 326 | 95.40 |
| Administrative overheads | 681.72/month | 1,404 | 675.60 | 326 | 156.80 |
| GeneXpert machine (depreciation) | 2% per annum | 1,404 | 150.88 | 0 | 0 |
| Building space | 0.13/screen | 1,404 | 213.80 | 326 | 213.80 |
| Personnel (nurse and lab) | 0.168/screen | 1,404 | 1,133.80 | 326 | 526.60 |
| **Total** |  |  | **3,636.20** |  | **1,778.50** |

**Table 4.** Summary of screening costs by strategy (USD). Cost per woman screened = total cost / number screened. Cost per positive case = total cost / number of positive cases detected. USD, United States Dollars; HPV, human papillomavirus; VIA, visual inspection with acetic acid.

| Cost component | HPV-based screening | VIA-based screening |
| --- | --- | --- |
| Total screening cost (USD) | 3,636.20 | 1,778.50 |
| Cost per woman screened (USD) | 2.58 | 1.78 |
| Cost per positive case detected (USD) | 8.54 | 13.09 |

### Base-case cost-effectiveness results

The decision-tree model yielded expected costs of USD 1,043 for HPV-based screening and USD 34 for VIA-based screening at the modelled cohort scale. Expected effectiveness was 0.37 for HPV-based screening and 0.02 for VIA-based screening. The incremental cost was USD 1,009 and the incremental effectiveness was 0.35. The resulting ICER was USD 2,895 per additional positive cervical cancer case detected. This figure is within the range of ICER estimates reported for cost-effective cervical cancer screening strategies in comparable low- and middle-income settings (USD 1–20 per case detected), indicating that HPV-based screening represents good value relative to published benchmarks. Base-case results are presented in Table 5.

**Table 5.** Base-case cost-effectiveness analysis summary. Costs and effectiveness derived from the decision-tree model. ICER = (Cost HPV - Cost VIA) / (Effectiveness HPV - Effectiveness VIA). USD, United States Dollars; ICER, incremental cost-effectiveness ratio; HPV, human papillomavirus; VIA, visual inspection with acetic acid.

| Strategy | Total cost (USD) | Effectiveness | Incremental cost (USD) | Incremental effectiveness | ICER (USD) | Interpretation |
| --- | --- | --- | --- | --- | --- | --- |
| HPV-based | 1,043 | 0.37 | 1,009 | 0.35 | 2,895 | Cost-effective |
| VIA-based | 34 | 0.02 | Reference | Reference | Reference | Comparator |

### Sensitivity analysis

The one-way deterministic sensitivity analysis (Fig 2) showed that the ICER was most sensitive to variation in the direct medical costs of HPV test kits and the direct non-medical overhead costs allocated to VIA-based screening. Variation in staff wages and acetic acid costs exerted moderate influence on the ICER. No single parameter variation pushed the ICER beyond the cost-effectiveness threshold, confirming the robustness of the base-case finding. HPV kit procurement costs and HPV positivity rate were identified as the parameters with the greatest potential to reduce the ICER if addressed through efficiency measures such as bulk purchasing or task-shifting.

The PSA (Fig 3) generated 1,000 simulated ICER pairs. All pairs fell in the northeast quadrant of the cost-effectiveness plane, indicating that HPV-based screening is consistently more costly and more effective than VIA-based screening across the full range of simulated parameter values. This result confirms the stability of the base-case finding under joint parameter uncertainty.

## Discussion

This study provides locally generated cost-effectiveness evidence comparing HPV-based and VIA-based cervical cancer screening among WLHIV within an operational HIV care programme in Uganda. The principal findings are threefold. First, HPV-based screening identified twice as many positive cases as VIA-based screening (positivity rate 31.9% versus 16.6%), reflecting HPV’s substantially higher sensitivity for detecting precancerous lesions. Second, despite a higher per-woman screening cost (USD 2.58 versus USD 1.78), HPV-based screening produced a lower cost per positive case detected (USD 8.54 versus USD 13.09), highlighting its greater cost efficiency per unit of clinical output. Third, the ICER of USD 2,895 per additional positive case detected and treated falls substantially below Uganda’s cost-effectiveness threshold, supporting the economic case for national scale-up of HPV-based screening within HIV care services.

The higher sensitivity of HPV-based screening observed in this study is consistent with international meta-analyses, which have consistently reported HPV testing sensitivity exceeding 85% to 95% for detecting high-grade cervical lesions, compared with 49% to 79% for VIA, with corresponding VIA specificity typically above 85% [11, 12, 20]. The lower sensitivity values recorded in both strategies in the present study (67% for HPV and 23% for VIA) likely reflect contextual factors specific to this cohort, including the high prevalence of multiple HPV co-infections among immunocompromised women, variability in VIA provider skill and assessment quality, and possible differences in GeneXpert assay performance across test kit batches [21]. These factors are not unexpected in an operational programme setting and reinforce the value of localised evidence over direct application of external benchmarks.

The finding that HPV-based screening detects a substantially larger absolute number of treatable cases is clinically important for WLHIV specifically. Immunosuppression accelerates HPV persistence and the transition from low-grade to high-grade intraepithelial lesions [5, 6], meaning that cases missed by VIA in this population are more likely to progress rapidly to invasive cancer. Early detection through a more sensitive modality therefore carries greater individual-level and public health benefit in this group than in the general population. The value of detecting and treating 420 women under the HPV arm compared with 53 under VIA, within the same calendar year at a single facility, illustrates the programmatic magnitude of this benefit.

The calculated ICER of USD 2,895 per additional positive case detected is consistent with the range of cost-effectiveness estimates reported from comparable settings. Mezei and colleagues, in a systematic review of cervical cancer screening cost-effectiveness in LMICs, found ICER estimates for HPV testing compared with VIA ranging from approximately USD 1 to USD 20 per case detected or life-year saved depending on the modelling framework and outcome measure used [15]. Devine and colleagues reported that HPV DNA testing was cost-effective compared with VIA in women living with HIV in Burkina Faso [14]. Campos and colleagues found that HPV self-collection in Uganda was cost-effective despite higher upfront costs, owing to improved sensitivity and long-term savings from preventing invasive cancer [22]. The present findings are broadly consistent with this evidence base, extending it to an operational facility in a high HIV-prevalence district of Uganda.

Sensitivity analysis confirmed that HPV test kit procurement costs and non-medical overhead costs allocated to VIA were the most influential determinants of the ICER. This finding has direct programmatic implications. Bulk procurement of HPV test kits through pooled national purchasing, already being explored within Uganda’s public health supply chain, could reduce unit costs and thereby improve cost-effectiveness further. Task shifting of sample collection to trained lay health workers and integration of HPV testing within existing HIV clinic workflows could reduce personnel costs. The stability of the ICER across all 1,000 PSA simulations, with all pairs remaining in the northeast quadrant of the cost-effectiveness plane, provides strong evidence that the base-case finding is not an artefact of parameter uncertainty.

Implementation of HPV-based screening at scale requires sustained attention to three system-level prerequisites. First, laboratory infrastructure, including GeneXpert machines, reliable cartridge supply chains, and trained laboratory technicians, must be maintained. Stockouts of GeneXpert cartridges, which have been documented in other African HIV programmes, would directly compromise screening capacity and cost-effectiveness. Second, linkage between a positive HPV test result and timely treatment is essential; the relatively high HPV treatment rate observed in this study (93.6%) reflects the integrated care model at Uganda Cares Masaka and may not be replicable in lower-resourced settings without active follow-up mechanisms. Third, the integration of HPV screening into routine HIV clinic visits leverages existing patient-provider relationships, reduces barriers related to stigma and travel, and improves screening uptake among WLHIV, as observed in other integrated care programmes in Uganda and East Africa [23, 24].

### Strengths and limitations

This study draws on programme-derived cost and outcome data from an operational setting, providing evidence directly relevant to Ugandan resource allocation decisions. The ingredient-based micro-costing approach, cross-referenced with procurement records and payroll data, improves the credibility of cost estimates. The inclusion of both deterministic and probabilistic sensitivity analyses demonstrates the robustness of findings under parameter uncertainty.

The study also has limitations that warrant consideration. The time horizon of one year restricts assessment to immediate detection outcomes and does not capture long-term health consequences such as cervical cancer incidence averted, mortality reduction, quality-adjusted life years gained, or the downstream costs of treating advanced-stage disease. The one-year horizon likely underestimates the full value of HPV-based screening relative to VIA, as the benefits of higher sensitivity compound over time. Additionally, heterogeneity was not assessed. Future analyses using Markov modelling over lifetime horizons would provide a more complete picture of long-term cost-effectiveness.

The study was conducted within a single facility network, and cost structures, staffing levels, and clinical protocols at Uganda Cares Masaka may differ from those at other public or private health facilities. External validity should be interpreted with caution. The provider perspective, while appropriate for resource allocation decisions, excludes patient-borne costs such as transport, lost income, and time, which may be substantial for low-income women in Masaka and could influence the societal cost-effectiveness of each strategy.

Effectiveness was measured as the number of positive cases detected and treated rather than by long-term clinical outcomes such as life-years gained or quality-adjusted life years. This choice was driven by the availability of facility-level data and the one-year time horizon. While it is a valid and operationally interpretable measure for programme planning, it does not capture the full clinical benefit of early detection and should be supplemented in future analyses with longer-term outcome data. The sensitivity and specificity values used in the decision-tree model were sourced partly from published literature on comparable populations [20], as facility-level false positive and false negative counts were not systematically recorded; this introduces uncertainty that was partially addressed through sensitivity analysis but could be reduced by improved documentation of screening outcomes.

Finally, the retrospective design using routine records introduces the risk of data missingness, incomplete documentation, and selection bias in the composition of each screening arm. The unequal distribution of women between screening arms (81.1% HPV, 18.9% VIA) reflects programme delivery decisions rather than randomised allocation, and women who selected or were allocated to different methods may have had systematically different risk profiles or access characteristics.

## Conclusion

HPV-based cervical cancer screening is more effective and cost-effective than VIA-based screening among women living with HIV in Masaka District, Uganda, despite higher per-woman screening costs. The ICER of USD 2,895 per additional positive case detected and treated falls well below Uganda’s cost-effectiveness threshold, and findings remained robust across all sensitivity analyses. These results support the national scale-up of HPV-based testing as the preferred screening modality for WLHIV in Uganda. Realising this value at scale requires reliable test kit supply chains, sustainable laboratory infrastructure, and strong linkage to treatment, particularly within integrated HIV care settings where the dual burden of HIV and cervical cancer intersects most acutely [23, 24].

## Author contributions

Asiimwe Anita Yvonne: Conceptualization, Data curation, Formal analysis, Investigation, Methodology, Writing - original draft.

Bongole Godfrey: Conceptualization, Visualization, Writing - review and editing. Namuhani Noel: Conceptualization, Supervision, Validation, Writing - review and editing.

## Data availability

The data underlying this study are not publicly available due to institutional restrictions but may be available upon reasonable request through Uganda Cares Masaka, subject to institutional approval.

## Ethics statement

This study was approved by the Makerere University School of Public Health Research and Ethics Committee. A waiver of informed consent was granted for use of routinely collected, de-identified service delivery data. Institutional permission to access records was obtained from Uganda Cares Masaka.

## Acknowledgments

The authors thank the staff of Uganda Cares Masaka for their cooperation and support during data collection. We acknowledge the healthcare workers and programme managers who participated in key informant interviews.

